# Class switch towards non-inflammatory IgG isotypes after repeated SARS-CoV-2 mRNA vaccination

**DOI:** 10.1101/2022.07.05.22277189

**Authors:** Pascal Irrgang, Juliane Gerling, Katharina Kocher, Dennis Lapuente, Philipp Steininger, Monika Wytopil, Simon Schäfer, Katharina Habenicht, Jahn Zhong, George Ssebyatika, Thomas Krey, Valeria Falcone, Christine Schülein, Antonia Sophia Peter, Krystelle Nganou-Makamdop, Hartmut Hengel, Jürgen Held, Christian Bogdan, Klaus Überla, Kilian Schober, Thomas H. Winkler, Matthias Tenbusch

## Abstract

Repeated mRNA vaccinations are an efficient tool to combat the SARS-CoV-2 pandemic. High levels of neutralizing SARS-CoV-2-antibodies are an important component of vaccine-induced immunity. Shortly after the first or second mRNA vaccine dose, the IgG response mainly consists of the pro-inflammatory isotypes IgG1 and IgG3 and is driven by T helper (Th) 1 cells. Here, we report that several months after the second vaccination, SARS-CoV-2-specific antibodies were increasingly composed of non-inflammatory IgG2 and particularly IgG4, which were further boosted by a third mRNA vaccination and/or SARS-CoV-2 variant breakthrough infections. While IgG antibodies were affinity matured and of high neutralization capacity, the switch in constant domains caused changes in fragment crystallizable (Fc)-receptor mediated effector functions, including a decreased capacity to facilitate phagocytosis. IgG4 induction was neither induced by Th2 cells nor observed after homologous or heterologous SARS-CoV-2 vaccination with adenoviral vectors. In addition, IgG2- and IgG4-producing memory B cells were phenotypically indistinguishable from IgG1- or IgG3-producing cells. Since Fc-mediated effector functions are critical for antiviral immunity, the described class switch towards non-inflammatory IgG isotypes, which otherwise rarely occurs after vaccination or viral infection, may have consequences for the choice and timing of vaccination regimens using mRNA vaccines.

## Introduction

During the ongoing pandemic of the Severe Acute Respiratory Syndrome Coronavirus 2 (SARS-CoV-2) with meanwhile over half a billion cases worldwide, new and efficient vaccines were developed with unprecedented speed and likely prevented millions of additional deaths^1, 2^. Alongside, this pandemic and the initiated vaccination campaigns have provided a unique opportunity to analyze *de novo* human immune responses to viral antigens after infection or immunization.

Among the first vaccine candidates, two mRNA vaccines (Comirnaty from Biontech/Pfizer and Spikevax from Moderna) were licensed in the United States and Europe, which were the first mRNA vaccines approved for use in humans. Both mRNA vaccines showed high efficacies of around 90% in preventing SARS-CoV-2 infections in clinical trials^3, 4^ and even in real-world scenarios^5–8^. The initially applied two-dose mRNA vaccination scheme induced robust antigen-specific immune responses. However, due to waning immunity and the emergence of immune escape variants of concern (VOC) like B1.1.529 (omicron), vaccine efficacies against SARS-CoV-2 infection dropped substantially^9–11^. Consequently, a third booster immunization with mRNA vaccines was recommended in various countries. Several studies confirmed that antibody responses after the third immunization were superior with regard to their neutralizing capacity against a broad variety of VOC compared to antibody responses measured after the initial two-dose regimen^12–14^. It was further shown that antibody avidity increased following mRNA booster vaccination, which was partly explained by prolonged germinal center (GC) activation and on-going B-cell maturation. SARS-CoV-2 vaccine-derived mRNA and spike protein was detected even several weeks after vaccination^15^. In-depth sequencing approaches on memory B cells documented somatic hypermutation (SHM) in GCs for up to six months, which resulted in broadening and diversification of the memory B cell repertoire, as well as effectiveness against VOC for which the vaccine was originally not designed ^13, 15–20^.

Activation-induced cytidine deaminase (AID) is the enzyme that catalyzes SHM in antibody variable (V) regions. It is expressed in GC B cells and also mediates class switch recombination (CSR) of constant (C) region genes^21, 22^. IgG3 encoded by the γ3 C-region is the most upstream (5’) Cγ-region in the immunoglobulin heavy chain gene locus on chromosome 14. Ongoing activity of AID can lead to switching towards more downstream Cγ regions, i.e. γ1, γ2 and γ4, which encode IgG1, IgG2 and IgG4^23, 24^. CSR is highly regulated during an immune response. The C-region to which a B cell switches is regulated by cytokines and B-cell activators at the level of transcription of unrearranged heavy chain constant genes^21^. However, the regulators for germline transcription of the γ2 and γ4 gene locus are not very well understood in humans. IL-4 in concert with IL-10 has been described to be involved in switching to IgG4^25^. Interestingly, distal IgG variants, in particular IgG2 and IgG4, were reported to mediate mostly non-inflammatory or even anti-inflammatory functions due to decreased Fc-mediated antibody effector functions including antibody-dependent cellular phagocytosis (ADCP), antibody-dependent cellular cytotoxicity (ADCC) or complement-dependent cytotoxicity (CDC)^23^.

Shortly after two doses of SARS-CoV-2-mRNA vaccination, IgG1 and IgG3 were the predominant IgG isotypes found in the vaccinated persons, whereas IgG2 responses were rare and IgG4 responses almost undetectable^26, 27^. This is in line with a Th1-biased cellular response seen after mRNA vaccination^28^. However, the longitudinal evolution of all four IgG isotypes (IgG1-IgG4) in response to mRNA vaccination – and particularly their long-term development after the second and the third (booster) dose – have not yet been analyzed.

Studying, two independent cohorts of vaccinated health-care workers, we report on a Th2-independent increase in anti-spike antibodies of the IgG4 isotype five to seven months after the second mRNA immunization. This response was further boosted by a third mRNA vaccination and/or by breakthrough infections with SARS-CoV-2 VOC. In line with the reported ongoing diversification of antibody responses, single-cell RNA sequencing (scRNA seq) of SARS-CoV-2 memory B cells revealed both avidity maturation and ongoing CSR resulting in IgG4 isotype generation. Although we confirmed increased antibody avidity and higher neutralization capacity against the recently emerged omicron VOC after the third vaccine dose, the switch towards distal IgG isotypes is accompanied by reduced fragment crystallizable (Fc) gamma receptor (FcγR)-mediated effector function such as ADCP. Considering the importance of Fc-effector functions in the control, prevention or treatment of viral infections, including SARS-CoV-2 in animal models^29–35^, this unusual vaccine response and IgG isotype switch might also affect subsequent SARS-CoV-2 infections in humans.

## Results

### Longitudinal monitoring of anti-spike antibody isotype responses

In a first cohort of 29 health care workers, we analyzed the antibody response after SARS-CoV-2 vaccination with three doses of Comirnaty (Table 1). The first two doses were given at an interval of three to four weeks and a further booster vaccination was applied about seven months after the second immunization. Using a previously established flow cytometry-based antibody assay^36^, anti-spike IgG responses were measured in sera taken ten days after each of the single vaccinations and 210 days after the second dose. In line with earlier reports^3, 4^, two mRNA immunizations induced robust IgG antibody response in all vaccinees with a median level of 697.5 µg/ml (Fig. 1A). This was followed by a significant decrease to 82.3 µg/ml during the following seven months, but antibody levels could be efficiently restored by a third immunization (median level of 699.7 µg/ml). Interestingly, the absolute amount of anti-spike IgG antibodies did not significantly differ 10 days after the second and the third dose. Neutralizing capacity was assessed in a surrogate virus neutralization assay confirming the dynamics of vaccine-induced antibody responses (Fig. 1B).

**Fig. 1:**
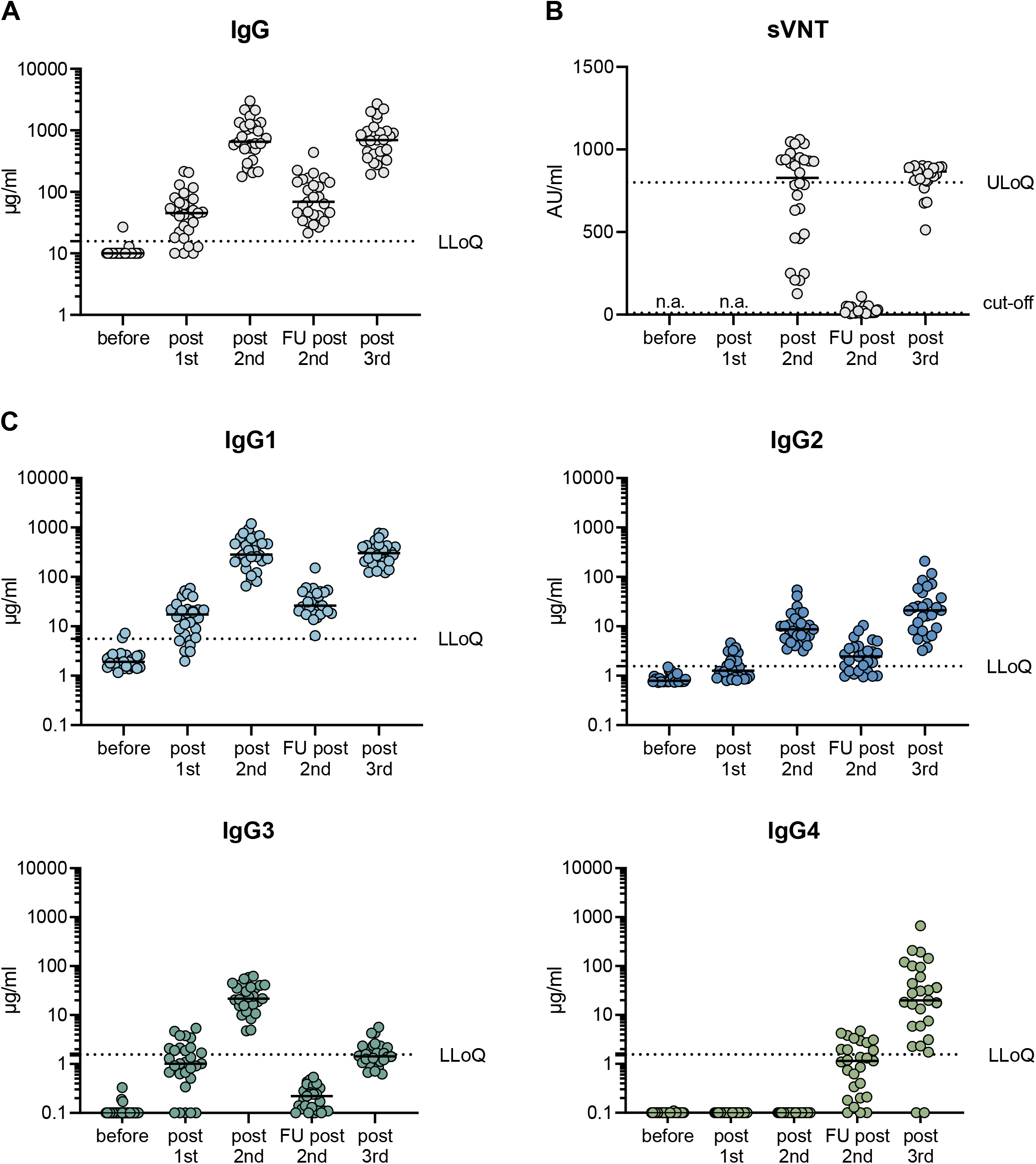
Longitudinal analyses of vaccine induced antibody response. 29 volunteers received three doses of the mRNA vaccine Comirnaty as detailed in Table 1. Serum samples were collected at a median of ten days after each vaccination (post 1^st^, post 2^nd^, post 3^rd^) and during a follow-up visit at 210 days after the second vaccination (FU 2^nd^). The total amount of spike-specific IgG was quantified by a flow cytometric assay using cell lines expressing full-length spike protein as targets (**A**). The neutralizing capacity was measured in a fully-automated surrogate virus neutralization assay. The latter was considered as positive within a linear range from 10-800 AU/ml. The dotted line indicates the cut-off value (**B**). The different IgG isotypes were quantified by flow cytometry using recombinant monoclonal receptor binding domain (RBD)-antibodies as a standard (**C**). The median MFI of three negative sera were used to set the background for each isotype. To visualize all sera in the graphs, all sera with MFI values below the background were set to 0.1 µg/ml. The lowest limit of quantification (LLoQ) is indicated in each graph by a dotted line and represents the lowest amount of the respective standard mAbs, which was detected (1.56 µg/ml for IgG2, IgG3 and IgG4; 5.6 µg/ml for IgG1). All individual values are depicted by open circles and the median is presented by the line. n.a.= not analyzed.

**Table 1:** Characteristics of study cohorts for longitudinal analyses after vaccination with three doses of Comirnaty

|  | Cohort 1<br>(CoVaAdapt) | Cohort 2 |
| --- | --- | --- |
| Vaccines | BNT162b2 mRNA (3x) | BNT162b2 mRNA (3x) |
| Number of volunteers | n = 29 | n=38 |
| Age in years, median (IQR)<br>[range] | 44 (31-52) [24-59] | 39.5 (28-53) [25-62] |
| Sex, n (%) Female | 16 (55%) | 27 (71%) |
| Sex, n (%) Male | 13 (45%) | 11 (29%) |
| Time intervals between<br>immunizations in days, median<br>(IQR) [range]<br>1<sup>st</sup> to 2<sup>nd</sup><br>2<sup>nd</sup> to 3<sup>rd</sup> | 23 (23-25) [22-28]<br>227 (216.5-229) [196-244] | 23 (22-28) [14-28]<br>207 (195-216) [170-251] |
| Time interval from immunization<br>to blood collection in days,<br>median (IQR) [range]<br>post 1<sup>st</sup>:<br>post 2<sup>nd</sup>:<br>Follow-up post 2<sup>nd</sup>:<br>post 3<sup>rd</sup> | 10 (9-10.5) [8-13]<br>10 (10-11) [9-11]<br>210 (209-210) [195-235]<br>10 (10-10) [9-12] | 14 (14-14) [7-18]<br>14 (14-17) [13-28]<br>165 (154-172) [147-194]<br>17 (14-23) [13-61] |

To study IgG isotypes, we generated recombinant monoclonal anti-receptor binding domain (RBD) antibodies composed of the four different constant heavy chains, but sharing the antigen binding Fab fragment. This allowed standardized quantification of the different anti-spike IgG isotypes in a multiplexed flow cytometric assay (see Methods). Ten days after two immunizations, anti-spike specific antibodies of the isotypes IgG1, IgG2 and IgG3 were readily detectable, whereas anti-S IgG4 antibodies were completely undetectable (Fig. 1C). IgG2 levels were markedly lower than IgG3 and IgG1 levels. Intriguingly, 210 days after the second immunization, the levels of spike-specific IgG4 antibodies exceeded the lower limit of quantification in the sera of about half of the vaccinees. The levels for all other isotypes dropped significantly as expected from the overall anti-S response. While IgG1 was still the most dominant isotype, IgG3 levels fell below the lower limit of quantification. After the third immunization, the amounts of IgG1 were elevated again and reached levels as measured shortly after the second vaccination. Notably, a marked increase in IgG4 antibody levels was observed after the booster immunization in nearly all vaccinees. In some individuals, IgG4 became the second most abundant isotype among the anti-S antibodies. Similarly, IgG2 levels increased after the third shot, whereas the elevation of IgG3 was marginal and only reached the levels seen early after the initial doses.

### Specificity of IgG4 induction through repeated mRNA vaccination

Having seen this increase of IgG4 antibodies late after the second immunization, we next explored whether this was specific for the (homologous) mRNA vaccination regimen. Therefore, we analyzed sera from a previously published cohort^37^, in which we compared the immunogenicity of homologous and heterologous vaccination regimens with Comirnaty and the adenoviral vector based vaccine ChAdOx1 (AZD1222, Vaxcevria) (see Suppl. Table 1). Five to six months after the second immunization, IgG1 was confirmed to be the dominant isotype after each of the three vaccination schedules (Fig. 2). Furthermore, vaccinees having received the homologous BNT-BNT or the heterologous Chad-BNT immunization had higher levels of anti-S antibodies as compared to recipients of two doses of Vaxcevria (Chad-Chad). More interestingly, spike-specific IgG4 antibodies were detectable in half of the sera of the BNT-BNT cohort (confirming the data obtained with cohort 1 in Fig.1), but only in one of the 51 sera from the other two vaccine cohorts (Fig.2).

**Fig. 2:**
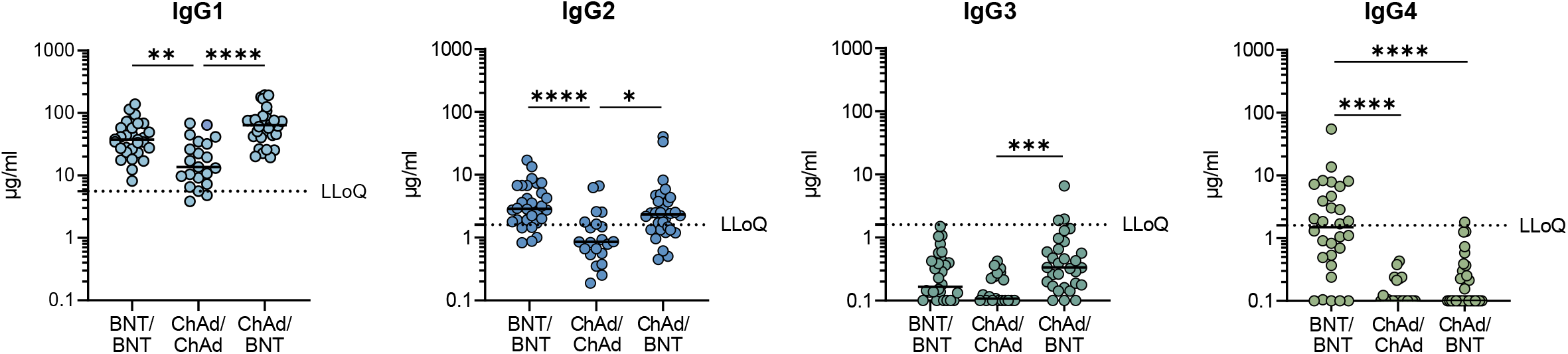
Comparison of IgG isotype specific antibody response. Anti-S specific antibody responses were analyzed in 81 individuals around five months after the second vaccination (Supplementary Table 1). 21 individuals had received two doses of the adenoviral vector vaccine, Vaxzevria, 30 had received an initial prime immunization with Vaxzevria followed by a boost with Comirnaty and an additional 30 were vaccinated twice with Comirnaty. The different IgG isotypes were quantified by flow cytometry. To visualize all sera tested in the graphs, sera with MFI values below the background were set to 0.1 µg/ml. The lowest limit of quantification (LLoQ) is indicated in each graph by a dotted line and represents the lowest detectable amount of the respective standard mAbs. (1.56 µg/ml for IgG2, IgG3 and IgG4; 5.6 µg/ml for IgG1). All individual values are depicted by open circles and the median is presented by the line. Kruskal-Wallis followed by Dunn’s multiple comparisons test was used for inter group statistics. *p < 0.05, **p < 0.01, ***p < 0.001, ****p < 0.0001.

### Th1 phenotype in spike-reactive T cells from individuals with high IgG4 levels

Since the induction of antigen-specific IgG4 antibodies could be a consequence of IL-4/Th2-driven cellular responses^38, 39^, peripheral blood mononuclear cells (PBMCs) of representative donors with low or high levels of IgG4 antibodies were collected 100 to 120 days after the third vaccination with Comirnaty and stimulated with spike peptide pools. Upon peptide stimulation, the percentage of activated CD4^+^ T cells (identified by the simultaneous expression of CD137 and CD154) increased as compared to unstimulated control cells (Fig. 3A). In individuals with low or high levels of IgG4 antibodies, similar amounts of spike-reactive CD4 T cells could be found (Fig. 3B). The majority of S-reactive CD4 T cells thereby produced IFN-γ and/or IL-2 and only a small fraction was positive for IL-4, confirming that S-reactive CD4 T cells maintained a Th1 phenotype even late after the third immunization (Fig. 3C). Thus, the occurrence of IgG4 after repeated SARS-CoV-2 mRNA vaccination appears to be a Th2-independent process.

**Fig. 3:**
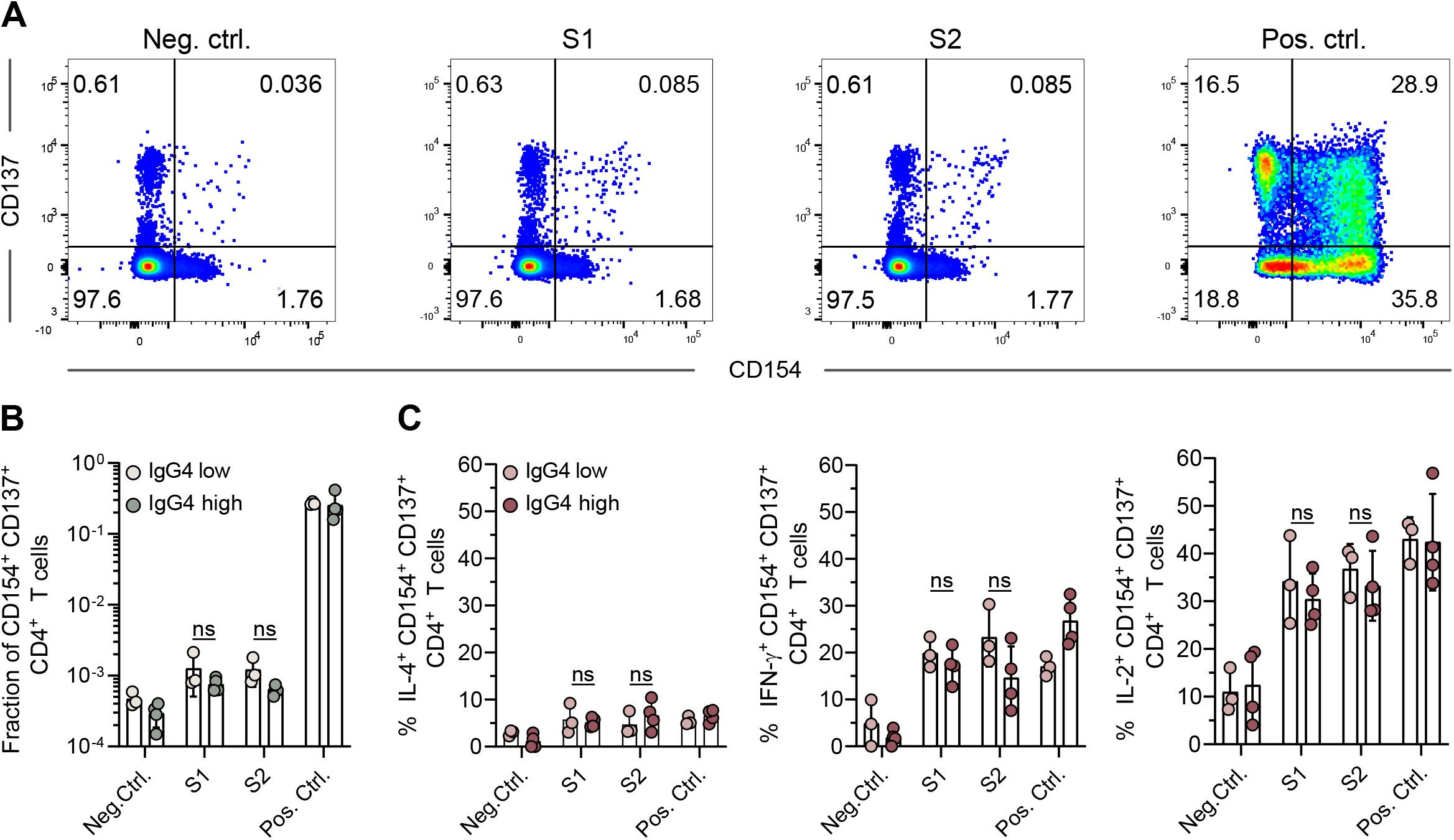
Characterization of spike-reactive CD4 T cells. Peripheral blood mononuclear cells of representative donors with low (n=3) or high (n=4) levels of IgG4 antibodies were collected 100-120 days after the third vaccination with Comirnaty and stimulated with S-reactive peptide pools S1 and S2. Unstimulated cells or cells stimulated with PMA/ionomycin were used as negative (neg. ctrl.) or positive controls (pos. ctrl.), respectively. Simultaneous staining for the activation markers CD137 and CD154 identified activated T cells among the CD4 T cells as shown for a representative donor (**A**). The fraction of activated T cells was compared between the donors according to their IgG4 status, IgG4^low^ vs IgG4^high^ (**B**). Within the activated T cell population, the percentage of IL-4, IL-2 and IFN-g producing cells was quantified and the comparison for IgG4^low^ vs IgG4^high^ is shown (**C**). No statistically significant differences could be observed between IgG4^low^ vs IgG4^high^ donors. Bars represent the median and circles represent individual vacinees.

### Transcriptomic profiling of phenotypes, somatic hypermutation and heavy chain isotypes of SARS-CoV-2-specific memory B cells

Both the antibody affinity maturation and the CSR of antibody constant (C) region genes (Fig. 4A) are driven by AID. Sequencing analyses of spike-specific memory B-cells have revealed a long period of antibody maturation after vaccination^16, 17, 20^. This might potentially come along with increased CSR towards distal IgG isotypes, which could result in the generation of IgG4-switched memory B-cells.

**Fig. 4:**
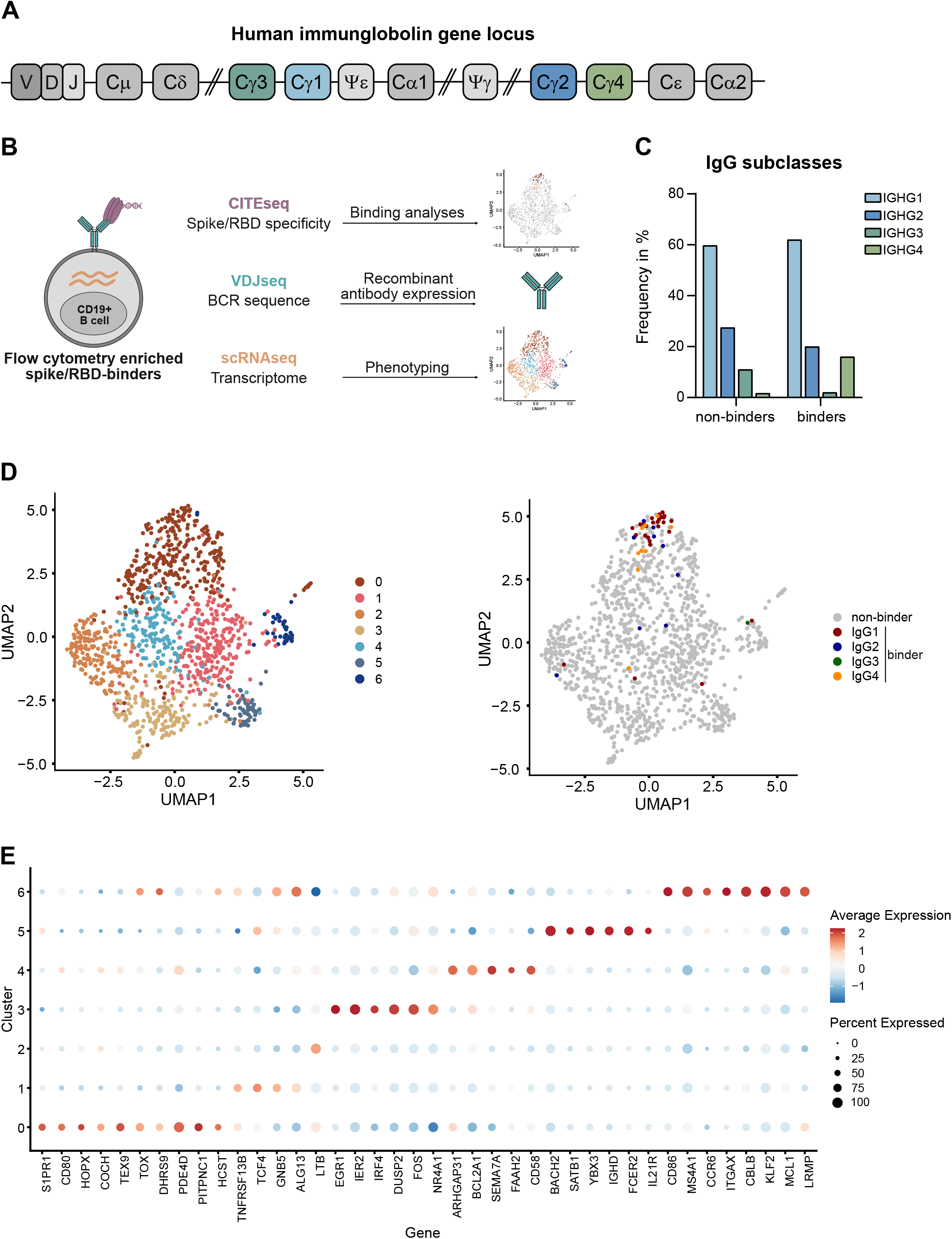
Single-cell RNA, CITE and VDJ sequencing of spike-binding memory B cells. (**A**) Schematic representation of the human immunoglobulin heavy chain gene locus. 5’ of each functional C-region (except Cδ) switch (S) regions are positioned directing class-switch recombination. Gene segments denoted by ψ resemble pseudogenes. (**B**) Experimental design for scRNA-seq data generation including CITE-seq and BCR-VDJseq. (**C**) Frequency distribution of IgG subclasses obtained from scRNA dataset. 4 individual donors from two timepoints (210 days post 2^nd^ and 10 days post 3^rd^ vaccination) were analyzed in parallel using hashtag-technology. Aggregate isotype frequencies of spike binders and non-binders are shown. (**D**) UMAP visualization of all CD19^+^ IgG^+^ sorted cells from all 4 donors and two timepoints that passed quality control. The cells are represented in seven main clusters (left). Spike binders were projected to the UMAP visualization with different colors depicting different isotypes (right). (**E**) Dot plot of cluster defining genes displaying the percentage of cells in a cluster, which expresses a certain gene (size) and the expression level (color). Red and blue colors indicate genes expressed above or below average, respectively.

To validate the presence of these cells within the memory B cell compartment and get insights into the diversity as well as transcriptional signatures of the respective memory cells, we performed scRNA seq from four selected donors 210 days after the second or 10 days after the third Comirnaty dose. For an unbiased comparison to other memory B cell populations in the same sample, we used cellular indexing of transcriptomes and epitopes (CITE) seq^40^ to label spike- and RBD-binding B cells, and enriched for spike-binding IgG+ B cells by flow cytometry prior to scRNA seq. (Fig. 4B). On aggregate, 16% of the sorted spike-specific memory B-cells from all four donors showed sequences encoding the IgG4 isotype (Fig. 4C). Frequencies of IgG4 anti-spike memory B cells for individual donors as determined through scRNA seq thereby mirrored serological IgG4 anti-spike levels (Suppl. Table 2). In contrast, IgG4 isotypes were barely detectable among non-binders (Fig. 4C). These results underscore the unusually high frequency of this type of vaccine-induced IgG isotype response.

To confirm the spike-specificity of the IgG4-producing B cell clones that were identified using CITE and scRNA seq, four recombinant monoclonal antibodies were cloned from the obtained B cell receptor (BCR) sequences, expressed in eukaryotic cells and tested for binding to RBD and full-length spike. All four monoclonal antibodies (mAbs) were able to bind to the spike protein, and one clone recognized RBD, which was in accordance with the flow cytometric sorting and the CITE seq results (Suppl. Fig. 1A).

Detailed analysis of the scRNA seq data revealed that irrespective of the IgG isotype almost all spike-binding memory B cells belonged to a cluster of cells (cluster 0) with a distinct expression profile that has not described for memory B cells before (Fig. 4D). Upregulated gene expression for antigen presentation (*CD80*) and a gene set found in switched memory B cells and memory CD8 T cells (*HOPX*, *TOX* and *COCH*)^41^ were found together with genes involved in signaling processes (*HCST/DAP10, PDE4D)* (Fig. 4E). Furthermore, we noted an upregulation of sphingosine 1-phosphate-receptor 1 (S1PR1) mRNA, which might indicate recent release from germinal centers^41^. Other distinct clusters among memory cells not associated with the spike-binding cells from the vaccinees included signatures for recently activated cells (cluster 3), naïve-like cells (cluster 5) and activated B lymphocytes with features of germinal center B cells (cluster 6, Fig. 4D, E).

To evaluate a potential correlation of ongoing SHM and CSR towards distal isotypes, we took a closer look at the SHM levels of spike-binding versus non-binding memory B cells. As described before^42^, we observed the highest numbers of somatic mutations in the V_H_ genes of the non-binding memory B cells using IgG4 (Suppl. Fig. 1B). In general, spike-binding memory B cells showed lower numbers of SHM when compared to total memory cells from the same isotype, including the IgG4 spike-binding cells (Suppl. Fig. 1B). This is compatible with the view that the mRNA vaccine response is still less diversified and/or affinity matured than the majority of all other memory B cells acquired over the lifespan of adults. Analysis of the limited numbers of available spike-binding memory B cells did not reveal significantly higher frequencies of somatic mutations in clones that use distal isotype sequences (Suppl. Fig. 1B).

In summary the scRNA seq data confirmed an unusually high frequency of IgG4 spike-binding memory B cells, which shared a specific transcriptomic profile with spike-binding IgG1 memory B cells.

### The IgG4 isotype does not prevail after repeated vaccination with tetanus toxoid or respiratory syncytial virus infection

Even after repeated immunizations or infections, IgG4 responses were only rarely observed. To corroborate this, we analyzed tetanus-specific antibody responses in 23 volunteers who had received several doses (2-16, median 6) of a tetanus toxoid (TT) vaccine (Suppl. Table 3). Sera were tested for TT-specific total IgG or IgG4 antibodies using an ELISA format. TT-specific IgG4 were detectable in 9 of 23 sera, albeit at very low levels, and no correlation was found with the number of vaccinations received (Suppl. Fig. 2A, B). Additionally, we tested ten individuals from our cohort 2 (Table 1) for the presence of antibodies against the respiratory syncytial virus (RSV), a respiratory pathogen that regularly causes re-infections in humans. While we found RSV-F protein-specific IgG1 antibodies in all tested sera, IgG4 was not detectable (Suppl. Fig. 2C). These findings support the notion that isotype switching to IgG4 is not a general consequence of repeated antigen exposures in form of vaccinations or infections.

### IgG4 occurrence correlates with increased avidity, but decreased antibody effector function in an independent cohort of SARS-CoV-2 mRNA vaccinees

To exclude any unrecognized bias or specific characteristics of the initially described cohort 1 (Table 1, Fig. 1), a second cohort of 38 volunteers was analyzed, who had received three doses of Comirnaty using a very similar vaccination schedule (Table 1, cohort 2). Sera taken shortly after the second or the third vaccination (2 to 5 weeks) showed a comparable anti-S antibody isotype distribution as described before (Fig. 1). Again, there was a substantial increase in IgG2 (3.5-fold) and IgG4 (38.6-fold) levels after the third immunization, whereas spike-specific IgG3 did not reach the levels seen after the second dose (Fig. 5A).

**Fig. 5:**
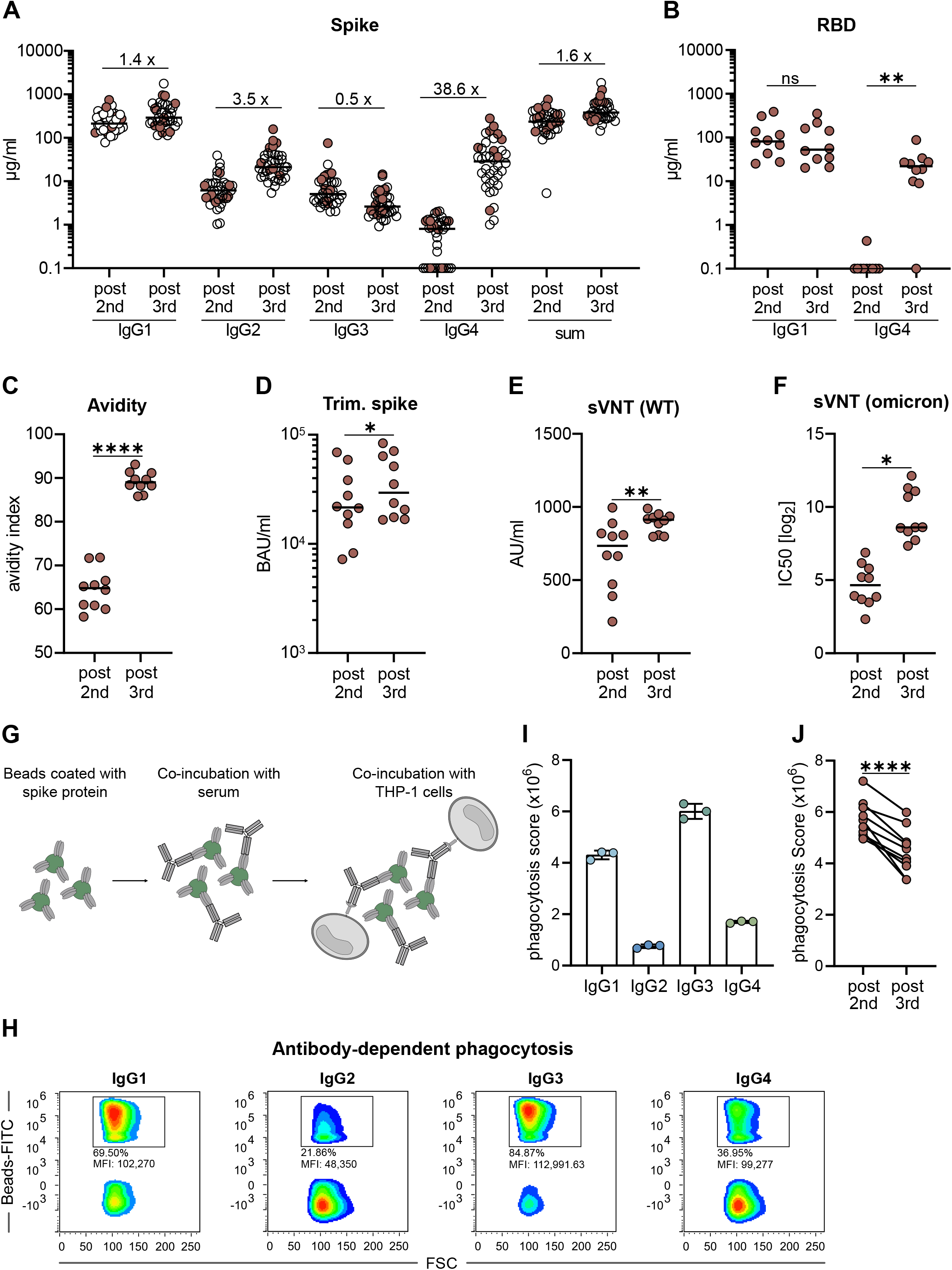
Comparison of functional antibody responses after two or three mRNA vaccinations. From a second cohort of 38 vaccinees having received three immunizations with Comirnaty (see Table 1, cohort 2), we selected ten persons to characterize the vaccine-induced antibody profile (indicated by filled circles). Paired serum samples collected after the second (post 2^nd^) or the third (post 3^rd^) vaccination were analyzed. The IgG isotype distribution measured in the flow cytometric assay and the sum of all IgG are shown for the whole cohort (**A**). The amounts of RBD-specific IgG1 and IgG4 (**B**) and the avidity (**C**) were determined by ELISA. For avidity measurements, sera were normalized according to total anti-S IgG levels and equal amounts of specific IgG were used. A fully automated CLIA assay was used to measure antibodies binding to trimeric spike protein. Antibody levels were quantified according to the WHO International Reference standard and given as BAU/ml (**D**). The neutralizing capacity was determined in a surrogate VNT against WT (**E**) and in a pseudotype VNT against the omicron VOC (**F**). Antibody-dependent phagocytosis by the monocytic THP-1 cell line was analyzed by using either monoclonal RBD antibodies of the different isotypes or the paired sera (**G-J**). Representative plots for IgG-mediated phagocytosis of spike-loaded microbeads by THP-1 cells are shown for the different isotypes (**H**) and the phagocytic scores for each monoclonal RBD antibody were quantified (**I**). The results from the paired sera are shown in (**J**). The phagocytosis score is calculated as follows: % of THP-1 bead-positive x mean fluorescence intensity of bead-positive. Circles represent individual sera and solid lines indicate the median. Statistical comparison for the two time points were done by a paired T-test. *p < 0.05, **p < 0.01, ***p < 0.001, ****p < 0.0001

Since the total amount of anti-spike IgG antibodies was only moderately (1.6-fold) elevated after the third compared to the second vaccine dose, we next investigated whether the increased proportion of IgG2 and IgG4 antibodies had functional consequences. To this end, paired sera from a representative sub-cohort of ten volunteers were analyzed. First, the presence of RBD-specific IgG1 and IgG4 antibodies was determined in an ELISA assay using Wuhan RBD as coating agent (Fig. 5B). RBD-specific IgG4 antibodies were significantly increased after the third vaccination, whereas the levels of RBD-binding IgG1 were not different at the two time points. Using sera that were normalized for their levels of total anti-S IgG, we found that the avidity was clearly increased after the third vaccination (Fig. 5C), which is in line with recent reports^12^. Furthermore, the capacities to bind trimeric spike protein (Fig. 5D) and to prevent soluble RBD binding to ACE2 (Fig. 5E), which both serve as surrogate markers for virus neutralization, were increased after the third dose. Accordingly, this translated in superior neutralization of lentiviral (LV) particles pseudotyped with spike proteins derived from the omicron VOC (Fig. 5F). In conclusion, repeated vaccination improved antibody effector functions mediated through the variable domain.

Since IgG2 and IgG4 are considered to have a lower potential to mediate FcγR-dependent secondary effector function, we also investigated Fc-mediated antibody effector function. To this end, we performed an ADCP assay with the monocytic THP-1 cell line^43^ (Fig. 5G). Using fluorescently labeled microbeads loaded with spike protein as targets and equal amounts of our recombinant monoclonal anti-RBD antibodies, we confirmed that IgG3 and IgG1 are more potent in inducing phagocytosis than IgG4 and IgG2 (Fig. 5H,I). Using FcγRIIA, FcγRIIB or FcγRIII-expressing reporter cells^44^, engagement of IgG2 and IgG4 results in reduced activation of the FcγRIIA, which was reported to be a key mediator of ADCP^43, 45^ (Suppl. Fig. 3). Consistent with this, sera taken after the third vaccination and normalized to the amount of anti-spike antibodies yielded significant lower phagocytic scores than sera from the same donors after two immunizations (Fig. 5J).

Together, these data show that spike protein-reactive IgG2 and IgG4 exhibit reduced Fc-mediated effector functions.

### Impact of breakthrough infections on vaccine-induced antibody responses

Finally, we asked whether subsequent infections with SARS-CoV-2 can also activate IgG4-switched memory B cells. We identified twelve persons from a study cohort of break-through infections (CoVaKo) who were vaccinated twice with SARS-CoV-2 mRNA vaccines and who experienced a breakthrough infection 25 to 257 days after the second mRNA vaccination (Suppl. Table 4). Infection was confirmed by PCR (2x Alpha, 7x Delta, 3x Omicron variant) and then serum samples were collected on the day of study inclusion (visit (V) 1, typically within the first week), two (V2) and four weeks (V4) after the PCR. In all individuals, we detected an anamnestic antibody response with an initial increase in spike-binding antibodies from V1 to V2 (Fig. 6). We also measured the kinetics of the different IgG isotypes in these samples (Fig. 6). All sera contained reasonable amounts of IgG1 and seven out of twelve had IgG2 antibodies above the lower limit of quantification at V1 (Fig. 6). In contrast, only two individuals’ V1 sera contained spike-specific IgG3 or IgG4 antibodies, respectively (Fig. 6). One additional patient developed IgG4 antibodies in response to the infection at V2. These three individuals with IgG4 antibodies above the lower limit of quantification at V1 experienced the infection 95, 201 or 257 days after the second vaccination, while in the other nine patients the infection took place between 25 and 78 days after the second mRNA shot. These latter nine patients without initial IgG4 levels also did not show a substantial boost of IgG4 antibodies during the three visits, while IgG1responses were boosted efficiently in all infected persons. Detectable levels of IgG2 were reached in all except one V4 sample, whereas the amounts of IgG3 remained at overall low levels. This supports our hypothesis that the switch to IgG4 is a consequence of ongoing GC maturation and that it takes several months until IgG4-switched memory B cells appear.

**Fig. 6:**
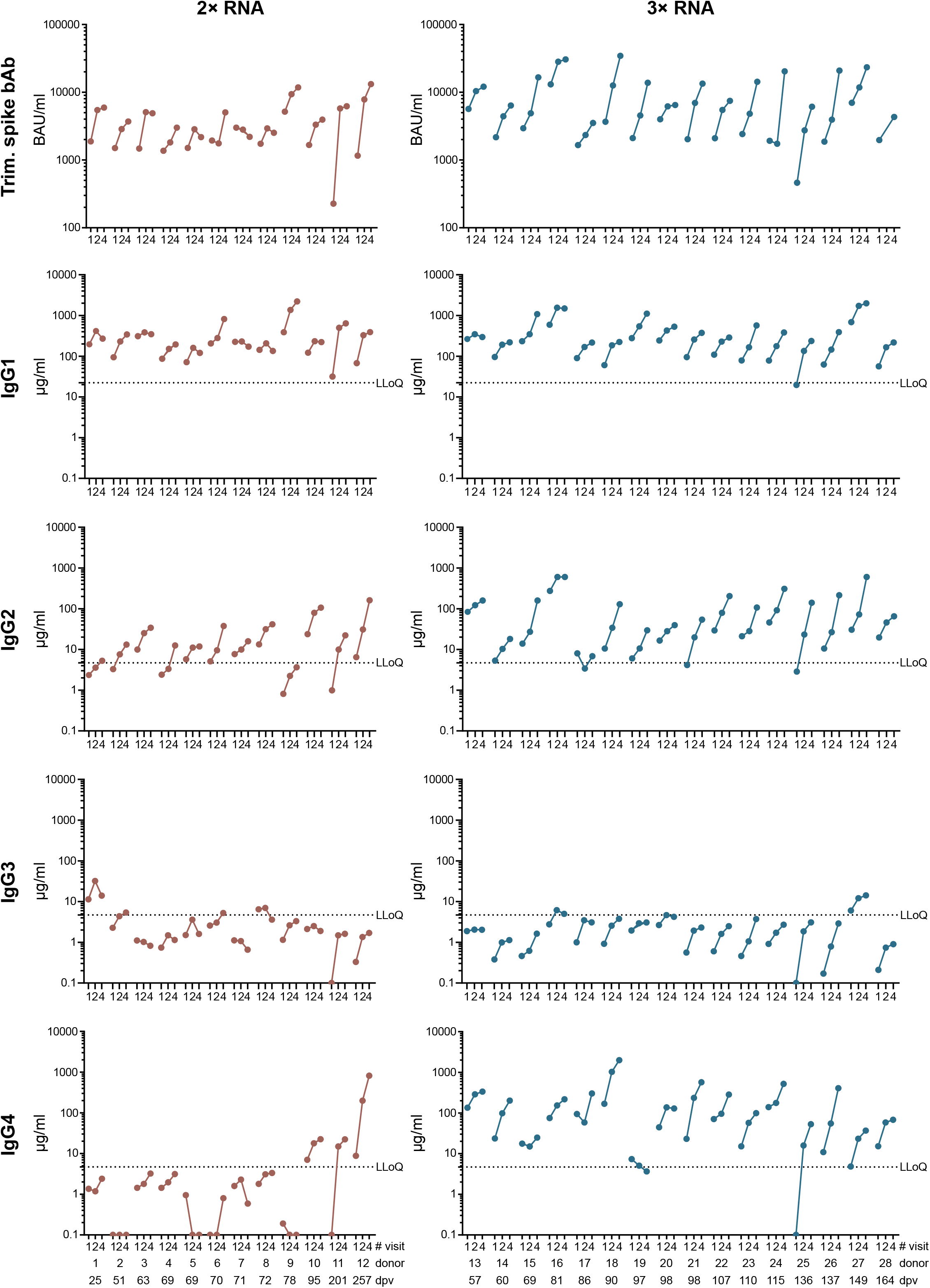
Anamnestic antibody responses after breakthrough infections. In individuals with PCR-confirmed SARS-CoV-2 breakthrough infection (see Suppl. Table 4), anamnestic antibody responses were quantified at the time point of study inclusion (V1, typically one week after PCR) as well as two (V2) and four weeks (V4) after the PCR result. Anamnestic responses in individuals experiencing infection after two (left panels) or three (right panels) immunizations were compared. Trimeric spike-binding IgG antibodies were analyzed in fully automated CLIA assay (LIAISON®SARS-CoV-2 trimericS IgG assay). Antibody levels were quantified according to the WHO International Reference standard and listed as BAU/ml. The levels of anti-S IgG1, IgG2, IgG3 and IgG4 were quantified by flow cytometry. The lowest limit of quantification (LLoQ) is indicated in each graph by a dotted line and represents the lowest detectable amount of the respective standard mAbs (4.68 µg/ml for IgG2, IgG3 and IgG4; 22.4 µg/ml for IgG1). Individual responses were indicated by dots with a connecting line.

In a second cohort of 16 patients, all participants had received three doses of mRNA vaccines prior to their breakthrough infection with the omicron variant. Samples were collected until V4 as described before (Suppl. Table 4). The time between booster shot and infection ranged from 57 to 164 days. Four of the 16 persons had a mixed immunization with two initial doses of Comirnaty and a booster with Spikevax, while the others received three doses of Comirnaty. In line with our data on the two previously described cohorts 1 and 2 (see Fig. 1; Fig. 5; Table 1), all except one of the triple vaccinated individuals had detectable amounts of anti-S serum IgG4 antibodies already at V1, although at levels varying between 5 µg/ml up to 168 µg/ml (Fig. 6). Throughout the observation time (V1 to V4), anti-S IgG4 levels further increased after breakthrough infection. In three individuals (donors 17, 18, 24) IgG4 was already the most dominant isotype of the anti-S antibodies at V1 and the isotype showed the highest proportional increase during the course of analysis. In the other individuals, the increases of IgG1 and IgG4 isotypes were comparable. This demonstrates that additional antigen contacts by infection can further drive the antibody response towards an IgG4 isotype.

## Discussion

In the present study we longitudinally tracked the antibody response in volunteers vaccinated with two or three doses of Comirnaty for a period of at least 8 months after the first vaccination. We made the unexpected observation of an mRNA vaccine-driven expansion of memory B cells expressing IgG4. The typical peak of the initial spike-specific antibody response shortly after the first two doses was followed by a strong decline over the next five to seven months, until the booster effect by the third mRNA vaccine dose became apparent. Interestingly, the total amounts of S-specific IgG antibodies were comparable at the peak after the second and shortly after the third vaccination, but there was clearly a different quality of the antibodies after the booster vaccination. In terms of antigen recognition, this was demonstrated by a higher avidity of the antibodies and a higher neutralizing capacity against the recent omicron VOC, which is fully in line with other recent studies^12–14^. The increased breadth with considerable diversification of the antibody repertoire is attributed to very long periods of antibody maturation after SARS-CoV-2 mRNA vaccination. B cell sequencing approaches have revealed the emergence of new S-reactive B cell clones even several months after the last vaccination^13, 15–19^. Interestingly, we found detectable levels of anti-S IgG4 antibodies in about half of the serum samples collected five to seven months after the second immunization, all of which did not show any IgG4 at earlier time points. For all other isotypes, a decline was seen in the same period. Moreover, after the third immunization, IgG4 levels sharply increased and became detectable in almost all vaccinees.

In line with the proposed ongoing GC reaction, the appearance of IgG4 antibodies might be a consequence of consecutive events of CSR and the maturation of IgG4 switched memory B cells. IgG3 antibodies were less efficiently boosted and did not reach the levels seen after the second dose. Considering the arrangement of the four γ heavy chain genes, which are ordered γ3-γ1-γ2-γ4 in the immunoglobulin gene complex on chromosome 14^24^, this would support the hypothesis of consecutive CSR from proximal IgG3 to distal IgG4^42, 46^. Interestingly, it is reported for the adult immune repertoire that CSR towards IgG2 or IgG4 is more frequently occuring from IgG1 B cells than from IgM/IgD cells^46^.

When we isolated spike-specific memory B cells from vaccinees ten days after the third vaccination, we confirmed by single-cell sequencing the presence of substantial numbers of S-reactive, IgG4 switched B cells, whereas IgG3-positive clones were hardly detectable. We cannot formally rule out *de novo* switching towards IgG4 immediately after booster vaccination. However, the presence of IgG4 antibodies in the sera at that time point, together with the rapid rise of IgG anti-spike serum antibodies, supports the idea of a reactivation of already present IgG4 memory B cells through the booster immunization.

Analysis of the frequencies of somatic hypermutations among spike-binders revealed lower levels as compared to the pool of all memory B cells for all isotypes in the adult individuals analyzed. This may be a sign for their relatively recent origin as compared to the much “older” memory pool archived throughout life^47^. Interestingly, our limited data regarding spike-binding IgG4 clones do not show higher mutation frequencies as compared to IgG1 clones. This is in line with very limited acquisition of new mutations in clones persisting over several months as described by Muecksch et al^13^. Similar frequencies of somatic hypermutations in spike-binding IgG1 and IgG4 clones suggest that both isotypes originate in parallel rather than consecutively. The indistinguishable gene expression profile of IgG1 and IgG4 clones further supports this view. However, the limited number of spike-specific B cells in our sample so far precluded analysis of clonally related pairs of IgG1 and IgG4 B cells, which makes definitive conclusions difficult. In any case, the mRNA vaccine itself and/or the narrow timing of the first two shots could be responsible for this effect.

In a cohort of breakthrough infections, the anamnestic IgG4 antibody response correlated with the time interval between immunization and infection. Individuals having experienced a breakthrough infection within the first 70 days after the second vaccination did not have substantial serum levels of anti-S IgG4 at their first visit, which also did not significantly increase during the following observation period. In contrast, anamnestic IgG4 responses were seen when breakthrough infections occurred later than 3 months after the second immunization, and were robustly detectable when the study participants had been vaccinated three times before infection. Although the number of subjects studied was limited and we did not stratify for potential confounding factors (e.g., the initial viral load, severity of disease or the VOC causing the infection), the presented data are consistent with the hypothesis of a slowly developing pool of IgG4 switched memory B cells after two doses of mRNA vaccination. Furthermore, we observed significantly higher IgG4 levels after two doses of mRNA compared to a heterologous immunization with a primary Vaxcevria vaccination followed by one dose of mRNA, although the total anti-S response was comparable. This argues against the hypothesis that repeated exposure to the spike protein itself triggers the unusual IgG4 response. Overall, our findings support the notion that the unusual vaccine-induced IgG4 response could be a specific feature of immunizations with mRNA vaccines or the relatively short interval between first and second immunization (three weeks) or both. It is currently not clear to what extent the mRNA vaccination or the short interval of immunizations are responsible for the observed long-lasting GC reactions.

Antigen-specific IgG4 antibody responses are generally thought to develop in response to Th2-mediated B cell activation or as a consequence of chronic antigen exposure^48, 49^. Parasite infections or allergic antigens are known for their potential to drive CD4^+^Th2 cells producing cytokines such like IL-4, IL-5 or IL-13, whereas most viral infections promote Th1-biased cellular responses^50^. Initial reports on the cellular response to the mRNA vaccine candidate BNT162b1 indicated a Th1 profile in the phase I/II clinical trial^28^ and also other reports did not identify Th2-biased immune response^51–53^. Against this background, Th2 responses appear to be a rather unlikely explanation for the robust and consistent IgG4 response we observed late after second and shortly after third immunization. Indeed, stimulating PBMCs from vaccinees showing high or low IgG4 levels with spike T cell epitopes did not reveal significant differences in the frequency of S-specific Th cells and rather suggested a Th1-biased CD4 response, based on the observed frequencies of IFNγ- and IL-4 producing cells. In contrast, a robust and persistent T follicular helper cell response for up to six month after mRNA vaccination has been characterized in draining lymph nodes by fine needle aspiration biopsis^20^, which is in line with a long ongoing GC reaction^15–17^. The underlying reason for this observation still requires clarification, but a prolonged presence of vaccine mRNA or antigen in the lymph node might be a potential explanation^15^.

Independent of the underlying mechanism, the induction of antiviral IgG4 antibodies is a very unusual phenomenon and raises important questions on its functional sequelae. Neutralizing antibodies preventing the initial binding of the viral particle to its specific cellular receptor are considered to be the most protective measure against SARS-CoV2 infections ^54^. The competitive binding would be mediated by the variable antigen binding site and does not rely much on the constant part of the Fc fragment. Indeed, we confirmed here previous reports on improved avidity and neutralizing potential of vaccine-induced antibodies after the third vaccination^12–14^. However, the vast number of breakthrough infections caused by the omicron variant indicate that the current vaccination does not confer sterile protection. Once infection is established, Fc-mediated effector functions become more relevant to clear viral infections. Systemic serology approaches have even revealed that different antibody functions can contribute to various degrees to protection dependent on the viral pathogen, as shown for Influenza Virus, RSV or SARS-CoV-2^55–58^. Passive immunization studies in animal models have further demonstrated that the degree of protection through the applied monoclonal antibodies depends on their IgG isotype^32–35^. In this regard, IgG4 is considered as an anti-inflammatory IgG with low potential to mediate Fc-dependent effector function such as ADCC or ADCP^23, 59^. High levels of antigen-specific IgG4 have been reported to correlate with successful allergen-specific immunotherapy by blocking IgE-mediated effects^60^. In addition, increasing levels of bee venom specific IgG4 have been detected in beekeepers over several bee keeping seasons and finally even became the dominant IgG isotype for the specific antigen, i.e. phospholipase A (PLA). Interestingly, the IgG4 response is characterized by a very slow kinetic and takes several months to appear, whereas PLA-specific IgG1 antibodies were already measurable at earlier time points, which resembles our findings in this study^61^. Furthermore, an increase in PLA-specific IgG4-switched B cells was observed in beekeepers during the season as well as in patients undergoing specific immunotherapies (SIT)^62^. For the control of viral infections not much is known regarding virus-specific IgG4 antibody responses. As shown here for RSV-specific IgG responses, IgG4 is hardly induced by acute respiratory viral infections even after repeated exposure. With the exception of measles-specific IgG4 antibodies induced by natural infection^63^, even chronic viral infections like HCMV do not trigger significant specific IgG4 antibodies ^63^.

There are very few reports on the induction of SARS-CoV-2 specific IgG4 after natural infection. The dominant isotypes were mostly IgG1 and IgG3^64–66^. Nevertheless, a Brazilian study during the early phase of the pandemic correlated an early onset and high levels of IgG4 antibodies with a more severe COVID-19 progression after SARS-CoV-2 infection, which might indicate a less effective antibody response^66^. However, in case of a primary immune response, the causality is difficult to address, since it is also possible that a more severe infection leads to an IgG4 response and not vice versa.

So far, only few studies on the role of vaccine-induced IgG4 responses against infectious diseases are available. In the field of HIV vaccine development, repeated protein immunization in the trial VAX003^67^ led to higher levels of HIV gp120-specific IgG2 and IgG4, whereas a prime-boost immunization with a canarypox vector (ALVAC) and the same protein vaccine in the RV144 trial^68^ resulted in higher HIV-specific IgG3 responses correlating with partial protection against HIV^69, 70^. Furthermore, the vaccine-elicited IgG3 Abs enhanced effector functions as ADCC and ADCP, but vaccine-induced IgG4 inhibited those functions^70^. Interestingly, for the malaria vaccine RTS,S an inverse correlation of serum opsonophagocytic activity (OPA) and protection against sporozoite challenge was reported^71^. Furthermore, changing the 0-1-2 month immunization schedule to a delayed fractional dose regimen with time intervals of 0-1-7 month and a reduced third dose, increased the vaccine efficacy of RTS,S which was paralleled by an increase in CSP-specific antibody avidity and higher somatic hypermutation frequency in CSP-specific B cells^72^. In a follow-up study, vaccinees showed reduced levels of OPA, which correlated with higher and more consistent IgG4 antibody responses after the delayed vaccine regimen^73^. This demonstrates that the quality and not only the quantity of vaccine-induced antibodies is important for vaccine-induced protection. Furthermore, the type of response can be directly influenced by changing the time intervals between the immunizations and the dosage of the booster vaccination.

In our study, antibody-mediated phagocytic activity was reduced in sera after the third immunization, in parallel to higher proportions of anti-S IgG4 antibodies. However, how these changes affect subsequent virus infections remains unclear. In a cohort of vaccinees with breakthrough infections, we did not obtain any evidence for an alteration of disease severity, which was mild in almost all of our cases. Larger cohorts with differential disease severities will be needed to address this aspect in the future. However, our results clearly demonstrate that a subsequent infection can further boost IgG4 antibody levels, with IgG4 becoming the most dominant IgG isotype among all anti-spike antibody isotypes in some individuals.

In conclusion, mRNA vaccines have proven enormous immunogenicity and efficacy, and have presumably saved millions of lives during this pandemic. A central reason for this is likely the potent GC reaction induced by mRNA vaccines, leading to affinity maturation and – probably more importantly – to diversification of a memory B cell pool, which can react to new antigen variants that the immune system has not yet encountered. However, as a collateral effect this immune response can also result in continuous CSR towards non-inflammatory IgG isotypes, which are capable to undermine some Fc-mediated effector functions. Our findings on an unusual, mRNA vaccine-induced antiviral IgG4 antibody response appearing late after secondary immunization clearly require additional investigation. Apart from further clarification of the precise underlying immunological mechanisms driving this response, it has to be evaluated how an IgG4-driven antibody response affects subsequent viral infections and booster vaccinations. This is not only relevant for potential future vaccine campaigns against SARS-CoV-2, but also for new mRNA-based vaccine developments against other pathogens.

## Methods

### Study cohorts

The cohorts are described in detail in Table 1 and Suppl. Tables 1-4. Ethics approval was granted by the local ethics committee in Erlangen (Az. 340_21B, Az. 46_21B, 350_20B and 235-18B).

### FACS-based antibody assay

The FACS-based antibody assay was adapted from a previously published assay to detect IgG, IgA and IgM antibodies^36^. Here, stably transduced HEK293T cells with doxycycline-dependent expression of SARS-CoV-2 spike protein derived from the Wuhan strain were used as target cells. In order to be able to quantify anti-S specific IgG1, IgG2, IgG3 and IgG4, we cloned and generated unique recombinant, monoclonal antibodies for each isotype with an identical RBD-recognizing variable region. The sequence for the BCR was obtained from spike-binding B cells isolated from an infected individual during the early phase of the pandemic. The recombinant mAbs were produced in HEK 293F cells. Purified mAbs were used to generate standard curves in each assay to allow quantification of absolute levels of the respective isotype in the tested sera. For a binding assay, spike protein expression was induced by doxycycline treatment for 48h, before 1×10^5^ cells were incubated with serum samples at various dilutions in 100 µl FACS-PBS (PBS with 0.5% BSA and 1 mM sodium azide) for 20 minutes at 4°C to bind to spike protein on the surface. After washing, bound S-specific antibodies of the different IgG isotypes were detected using the following antibodies: mouse anti-hIgG1-FITC (Sigma Aldrich, F0767), mouse anti-hIgG2-PE (SouthernBiotech, 9070-09), rabbit anti-hIgG3 (ThermoFisher, SA5-10204) followed by anti-rabbit-IgG-AF647 (ThermoFisher, A21443) and mouse anti-hIgG4-Biotin (SouthernBiotech, 9190-08) followed by Streptavidin-PB (ThermoFisher, S11222). After further washing, samples were measured on an AttuneNxt (ThermoFisher) and analyzed using FlowJo software (Tree Star Inc.). The median fluorescence intensities (MFI) correlate with the level of bound antibodies and standard curves for the different isotypes were generated by the statistical analysis software Graph Pad Prism9 (GraphPad Software, USA) using 4-Pl plotting. The median MFI of three negative sera were used to set the background for each isotype. To visualize all sera in the graphs, we set all sera with MFI values below the background to 0.1 µg/ml. The lowest limit of quantification is indicated in each graph and represents the lowest amount of the respective standard mAbs that were detected. (1.56 µg/ml for IgG2, IgG3 and IgG4; 5.6 µg/ml for IgG1 for a 1:100 dilution of the sera).

For the detection of RSV-specific antibodies, stably transduced HEK 293T cells with doxycycline inducible expression of RSV F-protein applying the same protocol as described above. The only exception was that there were no monoclonal standard Abs available. Thus, the MFI were plotted in the figure with different background values as stated in the figure legend.

### Automated measurement of trimeric spike antibodies

Sera collected from patients with breakthrough infections were analyzed for anti-spike antibodies using the fully automated LIAISON®SARS-CoV-2 trimericS IgG assay (DiaSorin, Saluggia, Italy) according to the manufactureŕs instructions. Antibody levels were quantified using the WHO International Reference standard and given as BAU/ml. An antibody level above 33.8 BAU/ml was considered as positive.

### Surrogate virus neutralization assay

For the detection of neutralizing antibodies, we ran the iFlash-2019-nCoV NAb assay on the iFlash-1800 CLIA Analyzer (YHLO Shenzhen, China) according to the manufacturer’s instructions. It detects antibodies that are able to compete with RBD binding to the SARS-CoV-2 receptor ACE2. According to the WHO standard, the neutralizing activity is given in AU/ml and the linear range for positive results is between 10 and 800 AU/ml.

### Antigen-specific antibody ELISA and avidity measurement

RBD-specific antibodies of the isotypes IgG1 and IgG4 were analyzed by ELISA. To this end, ELISA plates were coated with 100 ng of the RBD peptide (provided by Diarect GmbH, Freiburg) in 100 μl carbonate buffer (50 mM carbonate/bicarbonate, pH 9.6) per well over night at 4°C. Free binding sites were blocked with 5% skimmed milk in PBS-T (PBS containing 0.05% Tween-20) for 1h at RT. Serum samples were diluted 1:100 or 1:1,000 in 2% skimmed milk in PBS-T and incubated on the plate for one hour at RT. Recombinant monoclonal RBD antibodies were used as standard to allow the absolute quantification. After three washing steps with 200 μl PBS-T, anti-hIgG1-Biotin (dilution 1:2,000, MH1515, Thermo) or anti-hIgG4-Biotin (dilution 1:5,000, 9190-08, southernBiotech) were added for 1h at RT followed by an incubation with HRP-coupled streptavidin (1:2,000, ABIN376335, antibodies-online.com). Subsequently, the plates were washed seven times with PBS-T and after the addition of ECL solution, the signal was measured on a microplate luminometer (VICTOR X5, PerkinElmer).) and analyzed using PerkinElmer 2030 Manager software.

To estimate the avidity of the anti-RBD antibodies, sera were normalized for their anti-S antibody content and 100 ng of anti-S antibodies were used in the RBD avidity ELISA. This time, antigen-antibody complexes were incubated in the presence of 1.0 M ammonium thiocyanate or PBS as control for 30 min at room temperature. After washing to remove antibodies bound with low avidity, the ELISA was completed as described before. HRP-coupled anti-hIgG were used for the detection of bound antibodies. The relative avidity index was calculated as [IgG concentrations (NH4SCN) / IgG concentrations (PBS)] x 100, and is given in percent.

For the estimation of tetanus toxoid-specific antibodies, the same protocol as for the RBD ELISA was applied, but 0.5 µl/well of the inactivated TT vaccine (Tetanol®pur, Novartis) was used for coating. For the detection of TT-specific antibodies, either HRP-coupled anti-hIgG or anti-hIgG4-Biotin (dilution 1:5,000, 9190-08, southernBiotech) were used followed by incubation with HRP-coupled streptavidin.

### Pseudotype neutralization assay

Neutralization of recent Omicron variant was assessed with the help of spike-pseudotyped simian immunodeficiency virus particles as described before^74^. To produce pseudotyped reporter particles, HEK293T cells were transfected with the SIV-based self-inactivating vector encoding luciferase (pGAE-LucW), the SIV-based packaging plasmid (pAdSIV3) and the omicron spike-encoding plasmid

For the assessment of pseudotype neutralization, HEK293T-ACE2 cells were seeded at 2×10^4^ cells/well in a 96-well flat bottom plate. 24 h later, 60 µl of serial dilutions of the serum samples were incubated with 60 µl lentiviral particles for 1 h at 37°C. HEK293T cells were washed with PBS and the particle-sample mix was added to the cells. 48 h later, medium was discarded, and the cells washed twice with 200 µl PBS. Following 50 µl PBS and 25 µl ONE-Glo™ (Promega Corp, Madison, USA) was added and after 3 minutes the luciferase signal was assessed on a microplate luminometer (VICTOR X5, PerkinElmer) and analyzed using PerkinElmer 2030 Manager software. The reciprocal serum ID50 was determined with Prism GraphPad 9 (San Diego, California, USA) by application of the Sigmoidal 4PL function.

### FcγR activation reporter assay

The assay used for testing IgG-dependent activation of FcγRs is based on BW5147 reporter cells stably expressing chimeric FcγR-ζ chain receptors which stimulate mouse IL-2 production in the presence of Ag-IgG immune complexes, provided that the opsonizing IgG is able to crosslink the particular FcγR^44^. First, ELISA plates pre-coated with spike protein (kindly provided by InVivo Biotech Services GmbH, Hennigsdorf, Germany) were used to quantify the binding capacity of the different RBD-specific monoclonal antibodies. Ten-fold serial dilutions of the different mAbs (10 µg/ml to 0,001 µg/ml) were added to the plates and incubated 1h @ 37°C. After washing, plates were incubated with biotinylated anti-human IgG (1h, 37°C) followed by washing, addition of Streptavidin HRP (30 minutes at RT) and TMB substrate. Binding was quantified by measuring OD values at 450 nm and by calculating the area under the curve (AUC ELISA) To determine FcγR activation, serially diluted mAbs were incubated for 30 minutes at 37°C on spike pre-coated plates. Plates were thoroughly washed with RPMI 10% (v/v) to remove non-immune IgG and individual BW:FcγR-ζ reporter cells (expressing human CD16A, CD32A, CD32B CD64), were added to the spike-IgG complexes formed in the ELISA plate and incubated for 16 h at 37°C, 5% CO2. Afterwards, mouse IL-2 secretion was measured by anti-IL-2 ELISA, using purified rat anti-mouse IL-2 (BD-Pharmingen,) and biotin rat anti-mouse IL-2 (BD-Pharmingen, 1:500) ^44^. OD values were measured at 450 nm and the area under the curve was calculated (AUC IL-2). The IL-2 production in the supernatants was quantified as previously described^44^. The level of IL-2 as measure of the respective FcγR activation was first normalized to the total amount of spike binding antibodies, e.g. AUC IgG1 IL-2_CD16_/AUC IgG1 ELISA. Furthermore, the activation of the different FcγR by the IgG2, IgG3 and IgG4 isotypes were normalized to the respective activation by the IgG1 mAb to allow for inter-assay comparisons.

### Antibody-dependent phagocytosis assay

The phagocytosis assay was adapted from Ackerman et al^43^. Yellow-green fluorescent beads (FluoSpheres NeutrAvidin-labeled Microspheres, 1.0 µm; ThermoFisher, cat# F8776) were coated with biotinylated (Abcam, cat# ab201796) SARS-CoV-2 S1 Spike protein (Genscript, cat# Z03501) at a ratio of 100 ng protein per 5×10^8^ beads overnight at 4°C in FACS-buffer (PBS containing 0.5% bovine serum albumin and 1 nM sodium azide). After washing the beads with FACS-buffer, 5×10^6^ beads were seeded in 10 µl FACS-buffer per well in a 96-well plate. Monoclonal, spike-specific antibodies (1 ng mAb /well) or serum dilutions normalized to their spike-specific IgG concentration (1 ng anti-S /well) were added in a volume of 10 µl FACS-buffer. Sera were heat-inactivated at 56°C for 30 min. After an incubation of two hours at 37°C, 10^5^ THP-1 cells (ATTC TIB-202) were added to the beads in a volume of 100 µl RPMI 1640 supplemented with 10 % FCS, 2 mM L-Glutamine, and 1 % penicillin/streptomycin and the plates were incubated for 16 hours at 37°C. Cell-bead mixtures were washed two times with 180 µl PBS (400xg, 3 min) before treatment with 180 µl 0.25% Trypsin/0.02% EDTA for 10 minutes. After two additional washing steps with FACS-buffer, samples were resuspended in 200 µl FACS buffer and subjected to flow cytometry. Phagocytosis was assessed by first gating on THP-1 cells and then selecting cells that show yellow-green fluorescence of the phagocytosed beads. The phagocytosis score was calculated as follows: % of THP-1 bead-positive x mean fluorescence intensity of bead-positive. Data were acquired on an AttuneNxt (ThermoFisher) and analyzed using FlowJoTM software (Tree Star Inc.).

### Isolation and cultivation of peripheral blood mononuclear cell (PBMC)

PBMCs were isolated from citrate peripheral blood of vaccinated individuals by density gradient centrifugation using Biocoll^®^ separating solution, density 1.077 g/ml (Bio&Sell) and frozen in heat-inactivated FCS + 10% DMSO (Sigma-Aldrich) for liquid nitrogen storage. Thawed PBMCs were cultured in complete RPMI medium (RPMI 1640 medium (Thermo Fisher) supplemented with 10% heat-inactivated FCS, 50 µM ß-Mercaptoethanol, 0.05 mg/ml gentamicin, 1.192 g/l HEPES, 0.2 g/l L-glutamine, and 100 U/ml penicillin-streptomycin) at 37°C and 5% CO_2_.

### Intracellular cytokine staining

Cryopreserved PBMCs were thawed and rested overnight at 1×10^6^ cells/ml in complete RPMI medium. 10^6^ PBMCs were stimulated with 11aa overlapping 15-mer PepMixTM SARS-CoV-2 spike glycoprotein peptide pool (1 µg/ml), provided in two peptide sub-pools S1 and S2 (JPT), for 20 h at 37°C in the presence of 1 µl/ml GolgiPlug™ (BD Biosciences). For the unstimulated condition, PBMCs were cultured in complete RPMI medium and respective dilution of solvent DMSO. As a positive control, PBMCs were stimulated with 25 ng/ml phorbol myristate acetate (PMA) (Sigma-Aldrich) and 1 µg/ml ionomycin (Sigma-Aldrich). Following this incubation, all steps were performed at 4°C. PBMCs were washed twice with FACS buffer (PBS containing 0.5% BSA) and stained with ethidium-monoazide-bromide (EMA) (Thermo Fisher) for 15 minutes for live/dead discrimination. After two washing steps with FACS buffer, PBMCs were stained for surface markers CD8-eFluor450 (clone OKT8, Thermo Fisher, dilution 1:200) and CD4-APC-H7 (clone RPA-T4, Becton Dickinson, 1:100) for 20 minutes. Excess antibody was removed by two washing steps with FACS buffer followed by fixation/permeabilization using Cytofix/Cytoperm (BD Biosciences). PBMCs were washed twice with 1x Perm Wash buffer (BD Biosciences) and subsequently stained intracellularly for CD154-PerCP-Cy5.5 (clone 24-31, BioLegend 1:100), CD137-PE-Cy7 (clone 4B4-1, BioLegend 1:100), IL-2-APC (clone 5344.11, BD Biosciences, 1:20), IFN-γ-FITC (clone 25723.11, BD Biosciences, 1:10), and IL-4-PE (clone MP4-25D2, Becton Dickinson, 1:200) for 30 minutes. Following washing steps with first 1x Perm Wash buffer and then FACS buffer, PBMCs were filtered through a nylon mesh and acquired on a LSRFortessa^TM^ flow cytometer (BD Biosciences).

### Single B cell sequencing analysis

For labelling of spike-specific memory B cells a trimeric, prefusion stabilized spike protein^75^ fused to FusionRed was used. The spike ectodomain sequence was cloned into a pMT vector encoding a BiP signal peptide for efficient translocation, a FusionRed gene for FACS detection of spike-specific B cells and a C-terminal double Strep tag for efficient affinity purification of spike trimers. Protein expression was carried out in *Drosophila melanogaster* Schneider 2 cells as described before^76^. Trimeric Spike protein was purified by affinity chromatography from the supernatant using a StrepTactin Superflow column (IBA, Goettingen, Germany) followed by gel filtration chromatography using a Superose 6 10/300 Increase column (Cytiva).

For CITE-seq a unique molecular 5’ feature barcode was added to purified spike-FusionRed protein by the 5’ feature Barcode Antibody conjugation Kit – Lightning Link® (Abcam) exactly as described by the manufacturer. RBD was also labelled by a different feature barcode.

Live CD19^+^ IgG^+^ spike-FusionRed-binding memory B cells were sorted on a MoFlo Astrios Cell Sorter in the FACS-Core facility of the FAU. FusionRed-negative CD19^+^ IgG^+^ B cells were sorted in parallel and mixed with spike sorted B cells. FACS staining solution also contained barcoded hashtags for sample identifications (BioLegend) as well as barcoded RBD. Libraries for single-cell transcriptome sequencing and scBCR-sequencing were prepared using the, Chromium Single-Cell 5′ Library Gel Bead and Construction Kit, v2 Dual index Chemistry Kit and Chromium Single-Cell V(D)J BCR Enrichment Kit (10x Genomics, CA, USA). The scRNA-seq, CITE-seq and VDJ libraries were sequenced on the Illumina HiSeq-2500 platform in the NGS-Core facility of FAU. The scRNA-seq reads were aligned to the human reference genome GRCh38 (UCSC, CA, USA), after generation of cell-gene matrices via Cell Ranger v6.1.2 (10x Genomics). Cell Ranger v6.1.2 multi pipeline was applied for scRNA-seq and VDJ-seq analysis, while CITE seq Count v1.4.5 was applied to count the surface features^40^. Data analysis was further pursued with the R package Seurat v4.1.1^77^ under R v4.2.0. Cells with high mitochondrial gene content (>5%) were filtered out. Doublets and unlabeled samples were identified and filtered utilizing sample hashtags. For further analysis of antibody VDJ genes the IMGT/V-QUESTplatform was used^78^.

### Expression of recombinant antibodies

From the V_H_ and _VL_ sequences obtained by scRNA analysis, synthetic GeneBlocks were synthesized (IDTDNA) and cloned into expression vectors for human IgG1 and human Igk, essentially as described by Tiller et al.^79^, except that we used Gibson assembly for the cloning. Heavy and light chain plasmids were transfected into 293 cells and antibodies were harvested from the supernatants on day 3 after transfection.

## Data Availability

All data produced in the present study are available upon reasonable request to the authors

## Acknowledgments

The study was funded by the Bavarian State Ministry for Science and the Arts for the CoVaKo-2021 and For-COVID projects and the German Federal Ministry of Education and Science (BMBF) through the “Netzwerk Universitaetsmedizin”, project “COVIM” (to T.H.W. and H.H.). Further support was obtained from faculty COVID-19 fonds and the German Centre for Infection Research (DZIF). K.S. is supported by the BMBF (project 01KI2013) and the Else Kröner-Fresenius-Stiftung (project 2020_EKEA.127). K.K. is supported by the Deutsche Forschungsgemeinschaft (DFG) through the research training group RTG 2504 (project number: 401821119)

The funders had no influence on the study design and data interpretation. We thank Norbert Donhauser and Kirsten Fraedrich for excellent technical assistance.

