## Supplementary material for "Class switch towards non-inflammatory IgG isotypes after repeated SARS-CoV-2 mRNA vaccination"

**Supplementary Table 1**: Characteristics of study cohort comparing homologous vs. heterologous vaccination

| | Het-Imm (Erlangen)[37] | | |
|---|---|---|---|
| prime<br>boost | BNT162b2<br>BNT162b2 | ChAdOx1 nCoV-19<br>ChAdOx1 nCoV-19 | ChAdOx1 nCoV-19<br>BNT162b2 |
| Number of volunteers | n = 30 | n = 21 | n = 30 |
| Age in years, median (IQR) [range] | 47 (32-54)<br>[24-60] | 45 (40-55)<br>[31-59] | 46 (33-54)<br>[21-59] |
| Sex, n (%) Female | 15 (50 %) | 15 (71.4 %) | 15 (50 %) |
| Sex, n (%) Male | 15 (50 %) | 6 (28.6 %) | 15 (50 %) |
| Time intervals between immunizations in days, median (IQR) [range] | 23 (22-25)<br>[18-28] | 63 (63-63)<br>[63-63] | 63 (63-63)<br>[63-63] |
| Time interval from immunization to blood collection in days, median (IQR) [range] | 155 (153-164)<br>[148-177] | 142 (141-144)<br>[140-144] | 142 (142-144)<br>[141-144] |
| Overlap with Cohort 2 | 8 | 0 | 0 |

**Supplementary Table 2** Frequency of spike-binding memory B cells

| donor | anti-spike IgG4 210 days after 2nd (μg/ml) | Anti-spike IgG4 10 days after 3rd (μg/ml) | total number of spike-binding single-cells | Number (frequency) of spike-binding memory B cell | | | |
|---|---|---|---|---|---|---|---|
| | | | | γ1 | γ2 | γ3 | γ4 |
| 14 | 1.9 | 43.9 | 4 | 3 (75%) | 0 | 0 | 1(25%) |
| 15 | 0.7 | 2.3 | 13 | 9 (69%) | 4 (31%) | 0 | 0 |
| 22 | 4.2 | 143.2 | 27 | 17 (63%) | 6 (22%) | 1 (4%) | 3 (11%) |
| 31 | 3.1 | 665.5 | 6 | 2 (33%) | 0 | 0 | 4 (67%) |

### Supplementary Figure 1

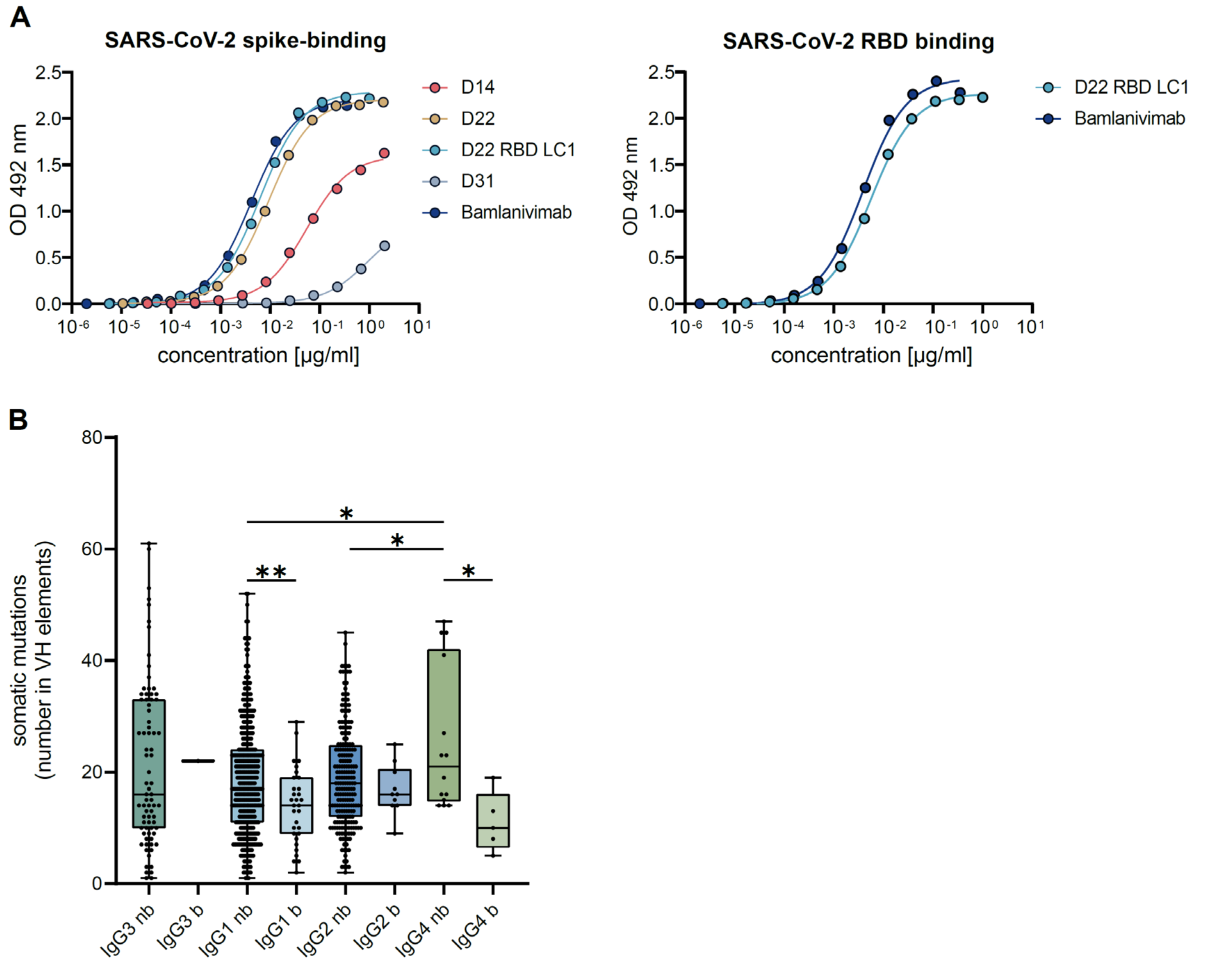

**Supplementary Figure 1:** Analysis of scRNA data.

(**A**) Characteristics of the produced recombinant antibodies derived from scRNA data from mRNA vaccinees. Heavy chain and light chain sequence information were extracted from 4 IgG4 spike binding cells obtained from scRNA. Left: ELISA binding curve to SARS-CoV-2 protein; right: ELISA binding curve to RBD peptide. The therapeutic SARS-CoV-2-specific monoclonal antibody Bamlanivimab was used as positive control. (**B**) Analysis of somatic hypermutations among spike binding and non-binding B cells as obtained from scRNA analysis. Numbers of mutated nucleotides for the VH gene segment are shown; t-Test, * $p<0.05$, ** $p<0.01$

**Supplementary Table 3**: Cohort with history of TT vaccinations

| Donor | Years since last dose | # of vaccine doses |
|---|---|---|
| U001 | 10 | 9 |
| U002 | 7 | 6 |
| U003 | 8 | 7 |
| U004 | 7 | 4 |
| U005 | 1 | 8 |
| U006 | 7 | 5 |
| U007 | 0.2 | 2 |
| U008 | 10 | 7 |
| U009 | 6 | 2 |
| U010 | 4 | 13 |
| U011 | 10 | 4 |
| U012 | 6 | 6 |
| U013 | 4.5 | 6 |
| U014 | 5 | 1 |
| U015 | 7 | 4 |
| U016 | 4 | 7 |
| U017 | 4 | 6 |
| U018 | 2 | 5 |
| U019 | 1 | 6 |
| U020 | 6 | 6 |
| U021 | 6 | 3 |
| U022 | 2 | 6 |
| U023 | 5 | 6 |

**Supplementary Figure 2**

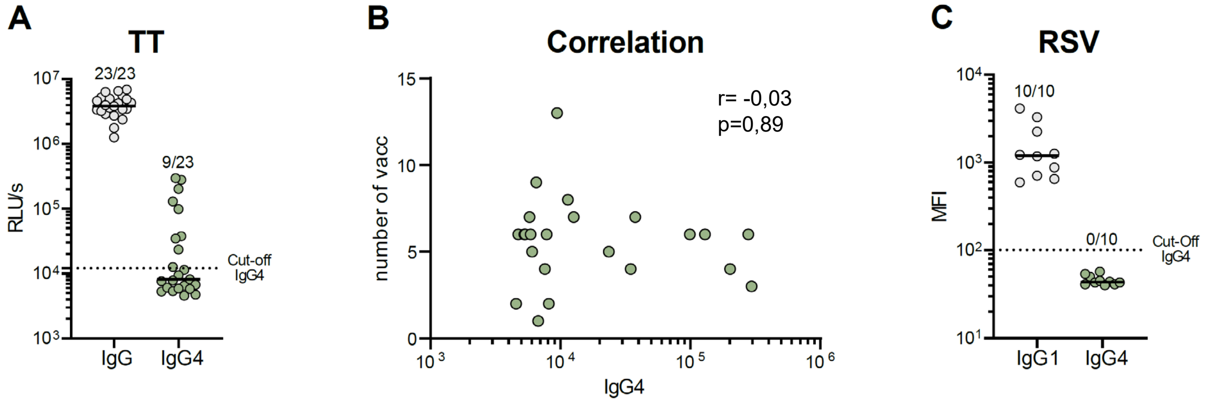

**Supplementary Figure 2**: IgG4 antibody response to tetanus vaccinations or RSV infections

In 23 individuals with a history of multiple vaccination against tetanus toxoid (see suppl. Table 3), TT-specific IgG and IgG4 antibodies were analyzed by ELISA (**A**). A non-parametric correlation were computed for the individuals IgG4 levels and the number of vaccine doses. The Spearman correlation coefficient and the p-value are shown (**B**). Using our FACS-based antibody assay with RSV-F protein expressing cells, RSV-specific IgG1 and IgG4 antibodies were measured in ten randomly selected sera of cohort 2 (**C**). Dots represent individual sera and lines the median. Number of positive sera are indicated for each analysis.

**Supplementary Figure 3**

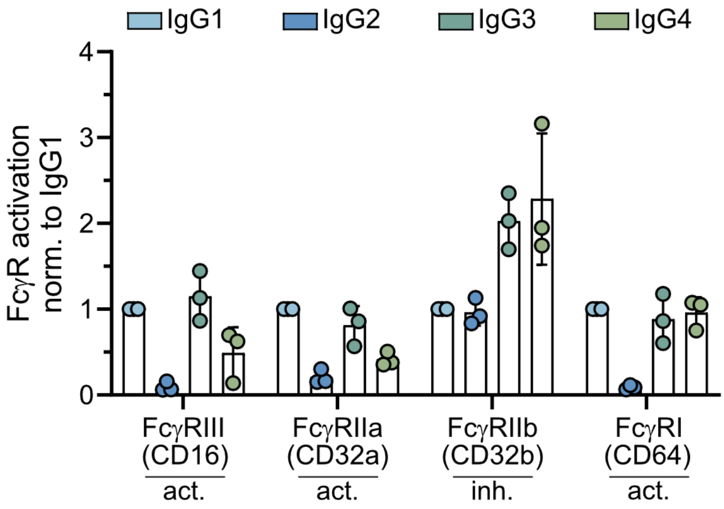

**Supplementary Figure3: FcγR reporter assay**

Using a panel of FcγR reporter cells assay, the monoclonal RBD antibodies (IgG1, IgG2, IgG3, IgG4) were analyzed for their potential to activate the different human FcR receptors CD16A, CD32A, CD32B and CD64. The level of IL-2 as measure of the respective FcγR activation was first normalized to the total amount of spike binding antibodies. Furthermore, the activation of the different FcγR by the IgG2, IgG3 and IgG4 isotypes were normalized to the respective activation by the IgG1 mAb. Dots represent each independent experiment (n=3).

**Supplementary Table 4**: Characteristics of cohort (CoVaKo study) with breakthrough infections after two or three mRNA vaccine immunizations

| Donor | gender | age group (years) | vaccines | VOC | distance last immunization to infection (days) |
|---|---|---|---|---|---|
| 1 | female | 50-59 | BB | Delta | 25 |
| 2 | female | 30-39 | BB | Delta | 51 |
| 3 | female | 50-59 | BB | Alpha | 63 |
| 4 | male | 30-39 | BB | Delta | 69 |
| 5 | female | 30-39 | BB | Delta | 69 |
| 6 | male | 20-29 | BB | Omicron BA.2 | 70 |
| 7 | male | 50-59 | BB | Delta | 71 |
| 8 | male | 20-29 | BB | Delta | 72 |
| 9 | female | 30-39 | BB | Omicron BA.2 | 78 |
| 10 | female | 50-59 | BB | Alpha | 95 |
| 11 | female | 30-39 | BB | Omicron BA.2 | 201 |
| 12 | female | 40-49 | BB | Delta | 257 |
| 13 | female | 30-39 | BBM | Omicron BA.1 | 57 |
| 14 | male | 15-19 | BBB | Omicron BA.2 | 60 |
| 15 | female | 30-39 | BBM | Omicron BA.1 | 69 |
| 16 | female | 30-39 | BBB | Omicron BA.1 | 81 |
| 17 | male | 40-49 | BBB | Omicron BA.1 | 86 |
| 18 | male | 30-39 | BBB | Omicron BA.1 | 90 |
| 19 | male | 60-69 | BBB | Omicron BA.1 | 97 |
| 20 | female | 20-29 | BBB | Omikron BA.2 | 98 |
| 21 | male | 20-29 | BBB | Omikron BA.2 | 98 |
| 22 | female | 20-29 | BBB | Omikron BA.2 | 107 |
| 23 | male | 40-49 | BBM | Omikron BA.2 | 110 |
| 24 | female | 60-69 | BBM | Omikron BA.2 | 115 |
| 25 | female | 40-49 | BBB | Omikron BA.2 | 136 |
| 26 | female | 30-39 | BBB | Omikron BA.2 | 137 |
| 27 | male | 50-59 | BBB | Omikron BA.2 | 149 |
| 28 | male | 50-59 | BBB | Omikron BA.2 | 164 |

B= Biontech/Pfizer; M= Moderna
